# Bacterial load slopes as biomarkers of tuberculosis therapy success, failure, and relapse

**DOI:** 10.1101/2020.05.03.20086579

**Authors:** Gesham Magombedze, Jotam G. Pasipanodya, Tawanda Gumbo

## Abstract

**Background:** Tuberculosis is expensive to treat, especially since therapy duration is at least six-months, and patients must be followed for up to two years in order to document relapse. There is an urgent need to discover biomarkers that are predictive of long-term treatment outcomes. Currently, tuberculosis programs use two-months sputum conversion for clinical decision making, while phase I clinical trials use extended [14 day] early bactericidal activity [EBA] to triage regimens. Our objective was to develop early treatment stage biomarkers that are predictive of long-term outcomes.

**Methods and Findings:** Data from 1,924 patients in the REMoxTB study was divided into [1] a derivation data-set of 318 patients on six-months standard therapy, [2] two sets of validation datasets comprised of 319 patients on six-months standard therapy, and 1,287 patients randomized to four-months experimental therapy. Sputum time-to-positivity [TTP] data was modeled using a system of ordinary differential equations that identified bacillary kill rates [termed *γ*-slopes], for fast-replicating bacteria [*γ_f_*] and for semi-dormant/non-replicating persistent bacteria [*γ_s_*], and to estimate time-to-extinction for all bacteria sub-populations in each patient. Time-to-extinction is used to predict the minimum therapy duration required to achieve cure. Using the derivation dataset, machine learning identified the *γ_s_* slope, calculated using first 8 weeks of therapy TTP data, as the highest ranked predictor for treatment outcomes. We then computed *γ_s_* slope thresholds that would reliably predict relapse-free cure for 2, 3, 4, and 6 months therapy duration regimens, and used these to create a diagnostic rule. In the first-validation dataset for six-months therapy duration, the *γ_s_*-derived decision rule demonstrated a sensitivity of 92% and a specificity of 89%; among patients with positive biomarker the relative risk [RR] of failure was 20.40 [95% confidence interval (CI): 7.17-58.08]. In comparison, two-month sputum culture conversion had a sensitivity of 33% and specificity of 71% [RR=1.20 (95% CI: 0.60-2.34)], while for extended-EBA sensitivity was 14% and specificity was 92% [RR=1.71 [95% CI: 0.73-3.48]. In the second validation dataset for four-months therapy duration, the *γ_s_*-derived diagnostic rule sensitivity was 81% while specificity was 87% for picking failure versus cure [RR=14.51 (95% CI: 8.33-25.41)]

**Conclusions:** The ability to predict treatment outcomes during the first eight-weeks of therapy could accelerate evaluation of novel regimens, development of new clinical trial designs, as well as allow personalization of therapy duration in routine treatment programs. Future research applying these diagnostic rules to different clinical trials data are required.

## INTRODUCTION

Tuberculosis [TB] is the most important infectious cause of death worldwide, accounting for 3% of all deaths; it killed one billion people over the last two centuries [1]. In both drug-susceptible TB and multidrug-resistant TB (MDR-TB) [2], therapy duration is 6 months, after which patients are followed for up to 18 months to document relapse. The large numbers of patients with TB [10 million/year], the long therapy duration, and the follow up period of up to 2 years, makes TB one of the most expensive diseases to treat. Thus, it is of crucial importance to identify TB treatment regimens that are equally as effective in drug-resistant TB as in drug-susceptible TB, to identify regimens that can shorten therapy duration, and to identify early biomarkers that obviate the need for 2-year follow up [1–11]. A closely related problem is the time it takes to evaluate and compare such new regimens in phase I-III clinical trials; they take decades to complete given the long follow-up time required to document relapse. Thus, biomarkers that obviate the need for the long follow up to document relapse, and that can be deployed immediately on a global scale at little cost, need to be urgently developed for both routine patient care and to accelerate the time-table of clinical trials.

The tools currently used to monitor TB treatment in the clinic and in clinical trials arose in the historical context of the microbiology technology of 50 years ago. In the late 1970s Jindani and Mitchison performed a 14-day treatment clinical study in East Africa [n=124 patients] that utilized solid agar-based *Mycobacterium tuberculosis (Mtb)* colony-forming unit [CFU]-derived kill rates defined by linear regression slopes to define early bactericidal activity [EBA], and the 14-day or extended-EBA to capture sterilizing activity, which are the basis of current phase I clinical trials [7, 8]. In 1993 Mitchison summarized results of seven clinical studies to propose the use of two-months sputum culture and smear as a surrogate of relapse; the two-month [eight-week] endpoint is now the basis of clinical decision-making in routine clinical care [3, 10–13]. Eight-week studies are also widely used as phase II studies to select TB regimens that go into the larger phase III studies in which long-term outcomes such as relapse, death, and cure are evaluated. However, the accuracy of these phase I/II studies in predicting hard clinical outcomes such as cure, therapy failure, and relapse, have been challenged [10–12, 14, 15]. In addition, more recent technological advances with semi-automated liquid cultures have demonstrated that the eight-week agar-based cultures may have been over-optimistic and are associated with substantial false-negative rates [16–19]. On the other hand, time-to-positivity [TTP] in the liquid cultures can be used in place of CFUs [20, 21]. The liquid culture technology is semi-automated and has been widely deployed across the world for routine clinical care as a diagnostic and for susceptibility testing. Here, we sought to identify mechanistic biomarkers (based on quantitative biology of the disease) that fulfill the definition of the US Food and Drug Administration BEST (Biomarkers, EndpointS, and other Tools) Resource, for use early during therapy to predict long-term hard clinical endpoints such as cure, therapy failure, and relapse[22, 23].

We have developed a mechanistic model to quantitatively explain the drug-regimen bacterial kill kinetics and dynamics of both fast-replicating and semi-dormant/non-replicating persistent [NRP] *Mtb* subpopulations in TB patients as reflected in sputum [24]. Here, we used serial sputum TTP-data from patients in the Rapid Evaluation of Moxifloxacin in Tuberculosis [REMoxTB] phase III clinical trial to identify the trajectory of these two bacterial sub-populations and to estimate time in which both *Mtb* bacteria subpopulations reach extinction (time-to-extinction) [24]. According to Burman, “The ability to prevent relapse is termed sterilizing activity because it is presumed to require killing nearly of all bacilli remaining after the initial phase of therapy” [9]. Restated, failure to reach extinction by the *Mtb* population in lung lesions is a required condition for therapy failure and relapse. Therefore, the time-to-extinction of all bacillary populations marks the required minimum duration of therapy in order to avoid relapse.

## MATERIALS AND METHODS

### Study design, data extraction and definitions

Our study design is reported in detail in **Figure. 1**. Briefly, we took data for bacteriologically confirmed TB patients that were enrolled in the REMoxTB clinical study [3]. In which patient sputum was cultured in the Mycobacteria Growth Indicator Tube [MGIT] to confirm bacteria viability. Since our aim was to develop a method agnostic of regimens used and drug-resistance status, patient data from the study [3] was used in our analyses regardless of drug-resistance status. Patients with majority of sputum samples that were contaminated or missing were excluded.

**Figure 1.**
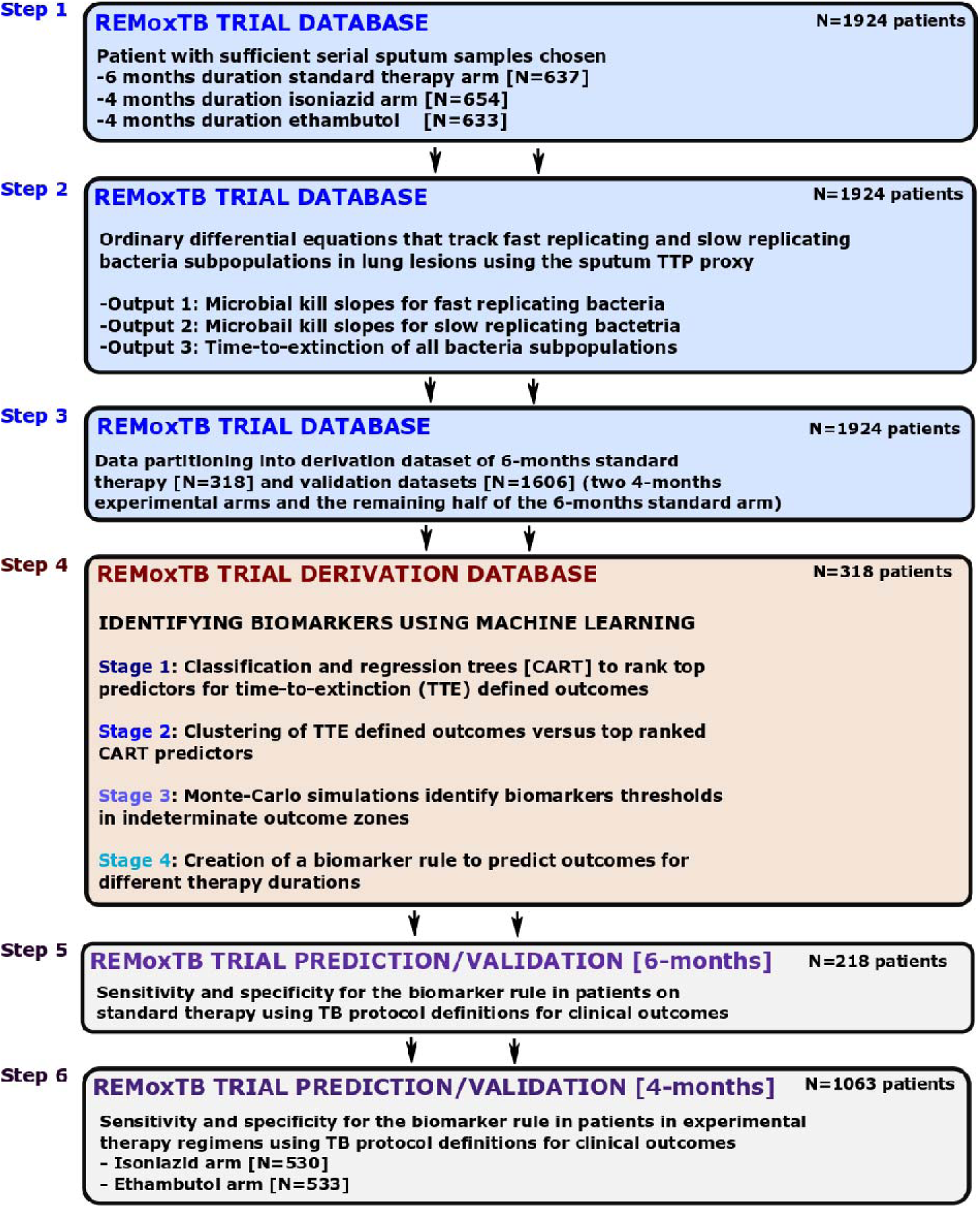
Biomarker Development Steps. **Step 1:** Patients without sufficient data points to derive bacterial kill slopes were removed. **Step 2:** The weekly sputum time-to-positivity data was then converted to colony forming units and the modeled using ordinary differential equations. **Step 3:** Data partitioning of 50% of patients in stanadrd of care six-months therapy as derivation data-set and the other 50% into valdiation dataset. All patients in experimental arm, administered over 4 months were assigned to validation datasets. **Step 4:** Four mathematical modeling and machine learning types of analysis in derivation dataset to [1] identify predictors of time-to-extinction [TTE] and [2] threshold values deliniating different TTE, and [3] design a diagnostic rule for different therapy durations. **Step 5:** Accuracy of diganostic rule/biomarker for six-months therapy duration in standard of care validation dataset using clinical definitions of outcome [relapse, cure]. **Step 6:** Accuracy of diganostic rule/biomarker for four-months therapy duration in two experimental arms in validation dataset using clinical definitions of relapse and cure.

Patient and microbial details, including therapy regimens and serial TTPs, were extracted from the CPTR website [http://www.cptrinitiative.org]. Time-to-extinction was defined as achieving a bacterial burden ≤10^ࢤ2^ colonies/mL, as mathematically justified in our prior work [24]. Microbiologic cure was defined as two negative sputum cultures without an intervening positive. Relapse was defined by the re-appearance of positive cultures in patients deemed cured at the end of therapy. Relapses were confirmed by 24-locus mycobacterial-interspersed-repetitive-unit analysis [24]. Failure to attain microbiologic cure at the end of therapy defined therapy failure, as per REMoxTB study protocol [24].

### Data partitioning

Patients on the standard TB therapy regimen were randomly partitioned into two subsets of equal size. The first set was designated as the model derivation set, while the remainder was assigned for use in model validation [validation data set]. To capture sufficient relapse events, only patients with at least two consecutive sputum samples during follow-up after treatment were used in model training and cross validation. Patients who received the experimental REMoxTB arms were used only in the validation dataset for sensitivity and specificity of predictors with 4 months therapy duration.

### Mathematical modeling for converting TTPs to CFUs

In order to convert TTPs to CFU/mL, we applied the formula:

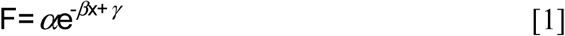

where *α* is 8.09 [95% confidence interval (CI) 6.64-9.96], β is 0.084 [95% CI: 0.08-0.087] and γ is 0.011[95% CI: −0.2 to 0.2], which we previously derived using more than 600 data point pairs from logarithmic phase growth and semi-dormant (or non-replicating phase) hollow fiber system model experiments [24]. Bacterial burden from these experiments were quantified using (i) solid agar culture for CFUs, and (ii) liquid medium in the MIGIT for TTP. The hollow fiber model is repetitively sampled for CFUs and TTPs for up to 56 days on therapy. Bowness and colleagues have found that as treatment progresses, the recovered Mtb grew more slowly in culture, so that a linear equation model [including only constants a, b, c] that remain unchanged during treatment would be incorrect by day 14, and instead a Gompertz model with a time parameter would be better [25]. While our formula is not a linear regression equation, we still wanted to find out if it was accurate at the start of therapy as at 56 days, in patients. Therefore, we applied formula/equation #1 to an independent clinical data set of patients on TB therapy, the vitamin A study in which we had weekly TTPs and CFUs in 56 patients as part of our morphism mapping between the hollow fiber system and patients on standard therapy [18, 24]. Results are shown in Figure S1, which shows that our formula remained accurate at 56 days as on day 0. Therefore, we employed equation #1 for toggling between CFU/mL and TTP.

### Mathematical Model

Our mathematical model, described in detail in the past [24], recapitulates events [i.e, *Mtb* burden] at site of infection, and, assumes two bacterial phenotypic populations: *B_f_*, fast replicating bacteria in log phase, which grows at rate *r_f_* and *B_s_*, non-replicating persisters which bacteria grow at rate *r_s_*, such that where *r_f_ >r_s_*, as observed by Canetti, McDermott et al, Sloan et al, Eum et al, and formalized by Mitchison [26–31]. Our assumption is that, in the lungs or at the site of infection, *Mtb* populations exhibit different physiological states, but share the same maximal bacterial burden, *K_max_* [32, 33]. The parameters *r_f_* and *r_s_* also measure of the reproductive or growth fitness, a measure of their virulence. The fast replication (log phase growth) *Mtb* grow at rate *r_f_* while the slow at rate *r_s_*. It has been shown that in TB patients, these bacteria subpopulations co-exist, however, in active TB disease, the population of bacteria in log-phase is dominant [26, 27, 29–31, 34].

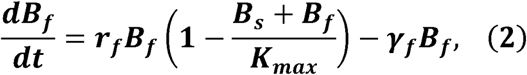

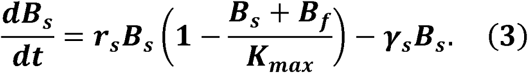

The model has flexibility to track the time evolution of both *Mtb* subpopulations simultaneously, under effect of treatment with different combination regimens. In relation to assessing new surrogate markers or biomarkers for predicting TB treatment outcomes, the model has two sets of quantifiable parameters (i) *r_f_, r_s_* and *K_max_ (Mtb* growth parameters) and (ii) *γ_s_* and *γ_f_* (drug-regimen based microbial kill slopes), that are linked to disease pathogenesis, and therefore has the ability to predict disease outcomes independent of a specific TB therapy regimen. Further mathematical details and assumptions of the model are shown in supplementary methods.

### ODE-based model to data fitting

First, all patient TTP longitudinal observations were converted to CFU values using equation 1. Then the data was fit to the system of ODEs (Equations 2 and 3). We implemented the Markov chain Monte Carlo (MCMC) method in R [22, 24, 35] to estimate the drug kill parameters using 50, 000 runs of the chain. A Gaussian log-likelihood was used to generate posterior distributions for parameters assuming uniform distribution for the priors. Model to data fitting was done to estimate the *γ_f_* and *γ_s_* slopes. The estimates were derived as medians of the MCMC posterior distribution, the uncertainty was given by 95% credible intervals (CrIs) calculated from the 2.5% and 97.5% quantiles of the MCMC parameter posterior distribution. MCMC convergence was assessed visually and by using the chain convergence diagnostic tools in the R coda package.

### Identification of biomarkers predicting outcomes in derivation dataset

Identification of biomarkers that best predicted therapy outcomes was carried out using classification and regression criteria [CART] of Breiman et al [36]. Using the derivation dataset, we examined all demographic, clinical, and radiological factors, as well as model-derived *γ_s_* and *γ_f_* slopes and the initial bacterial burdens [*B*(0)], as potential predictors of outcome. Outcome was defined as either therapy success at end of therapy, or therapy failure (failure at the end of treatment or relapse), or relapse. The steps we followed were implemented by two independent investigators in R (Rpart) and Salford software, and have been described in detail in the past [37].

First, CART analysis was used to identify and rank the top predictors of therapy failure and relapse. Second, we used clustering to characterize the relationship of the top predictors for each specific treatment outcome, and also identified the statistical association [38]. TTP trajectories were clustered using the K-means algorithm implemented in the KML-package in R [38]. The 6-month TTP data for each cluster was reduced to derive (i) the 4-month slopes [using the first 4 months accrued data] and (ii) then 2-month slopes [based on the first eight-week accrued data]. The model was fitted to data for each separate cluster and their respective reduced subsets.

Finally, we utilized Markov chain Monte Carlo simulations of time-to-extinction in tandem with CART to identify slope thresholds and initial bacterial burden that best classified relapses and therapy failure [35].

### Mathematical simulations for indeterminate data zones

We computed 10,000 bacteria trajectories to simulate different treatment outcomes. The initial bacterial burdens based on the range in derivation data set of between 3-7 log_10_ CFU/mL and *γ*-slopes between 0.05 to 0.5 log_10_ CFUs/day, were varied simultaneously, with the rest of all model parameters held constant. TTE for each separate trajectory was computed. The TTE values define the transcritical bifurcation points that explains when the Mtb NRP stable state switches to extinction. Regions of time within which bacteria subpopulations would go extinct were constructed and partitioned to reflect the expected clinical treatment duration intervals.

### Sensitivity analysis for treatment duration

Monte-Carlo experiments were carried out to identify changes in *γ_s_* values that resulted in treatment duration shortening (2 and 4 months) and those that led to prolonged treatment duration (7, 8 and 9 months). Magnitudes that correspond to these treatment end-points were determined relative to different categories of patient initial bacterial load, (i) high (>5·0 log_10_ CFU/mL), (ii) medium (3·5-5·0 log_10_ CFU/mL) and (iii) low (<3·5 log_10_ CFU/mL). These bounds were selected to toggle between CART discrete bounds and sweep across continuous patient CFU burdens to examine effect of different slope magnitudes on outcome for the defined therapy durations.

### Validation of identified biomarkers

Individual patient TTP trajectories were fitted to the model to identify the corresponding *γ_s_* and *γ_f_* in the validation datasets. The accuracy, sensitivity, and specificity of biomarkers derived in the derivation dataset were calculated using the validation dataset for cure, relapse, or therapy failure, for 6 and 4-months duration of therapy. The definitions for cure, relapse, and therapy failure used were those defined by the REMoxTB clinical trial protocol [3]. We used the standard statistical and clinical definitions for sensitivity, specificity, accuracy, and the number needed to diagnose failure and relapse [39, 40].

### Statistical analysis

Mean values between groups were compared using Student’s t-test or analysis of variance (ANOVA) F-test, while the Mann-Whitney test was used for proportions and compare medians from distributions of the fast and slow slopes derived at 2-months, 4-months and 6-months accrued TTP data. Spearman’s correlations were used to examine correlation while un-weighted Cohen’ Kappa coefficients examined agreements of clinical outcomes derived from REMoxTB study definition versus those derived from the model based on time-to-extinction. All analyses were performed with packages in R.

## RESULTS

### Clinical and laboratory characteristics in derivation and validation datasets

First, REMoxTB clinical trial patients who had insufficient serial sputum samples were removed, leaving 637 [33%] patients randomized to the standard therapy arm, 654 [34%] randomized to the isoniazid arm, and 633 [33%] randomized to the ethambutol arm [**Figure 1**]. This was followed by converting the 1,924 patients TTP-series to CFU/mL using equation 1, before modeling the data with a set of ODEs 2 and 3, to describe trajectories of *Mtb* CFU/mL with time [i.e., slopes]. We identified ODE-model parameter estimates using 8-week [2-months]-, 4-months-, and 6-months accrued TTP-derived data for all 1,924 patients. The model parameter estimates are shown in **Table S1**. We termed the *Mtb* kill rates *γ*-slopes, where *γ_f_* is the slope for fast-replicating *Mtb* and *γ_s_* is the slope for semi-dormant/NRP *Mtb*. The model was also used to calculate the time-to-extinction of the total *Mtb* population for each patient, with results shown in **Figure 2**.

**Fig 2.**
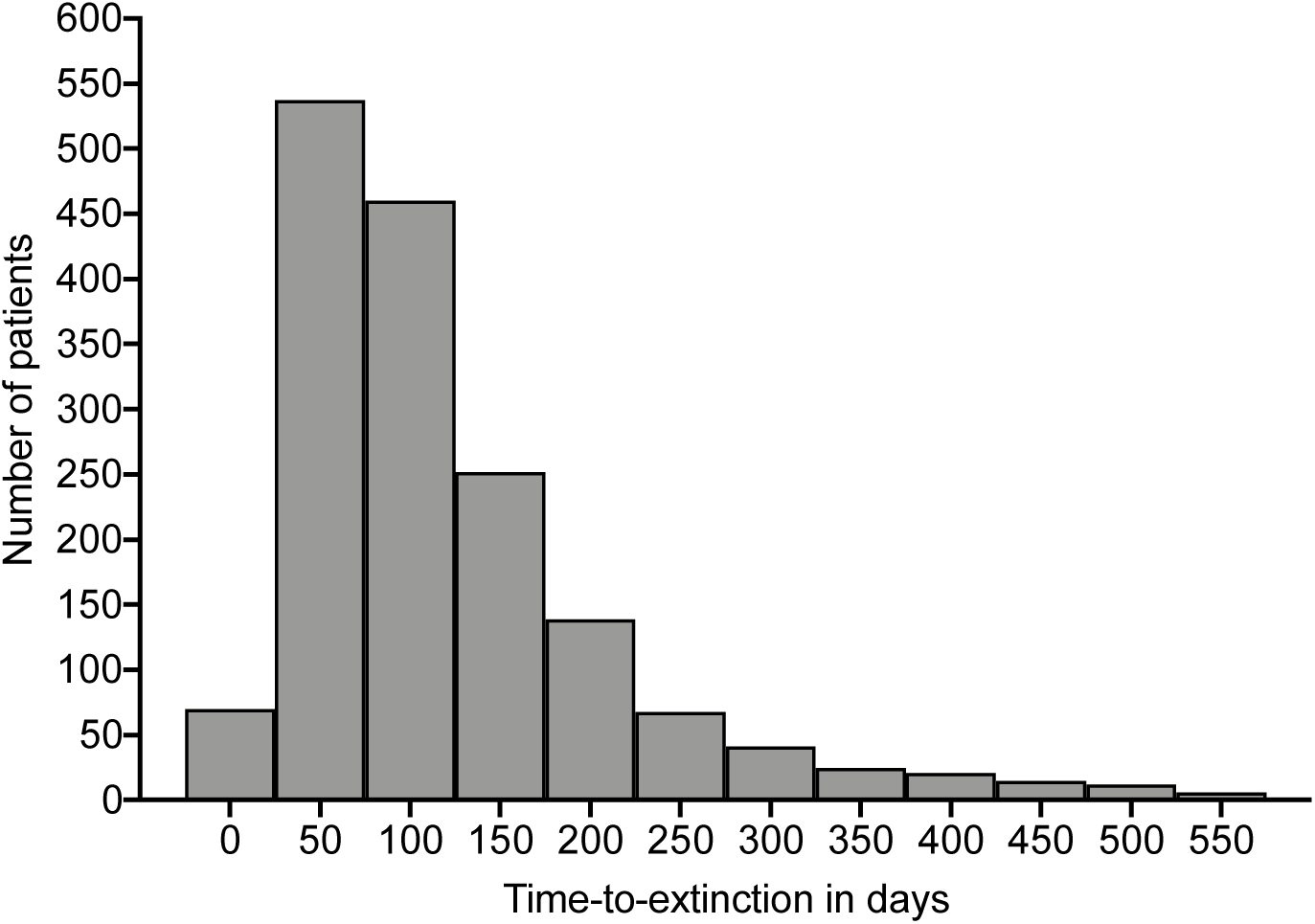
Distribution of time-to-extinction for all 1,924 patients. Shown is the data for all the 1,646 patients who achieved bacillary population extinction; bacilli in the remainder of patients did not reach extinction, so the time is at infinity. The mean time to extinction and 95% confidence intervals were 122.4 [117.9 to 126.9] days.

### Data partitioning into derivation and validation datasets

We separated the 1,924 patients’ data into derivation and validation datasets, shown in **Table 1**. The derivation dataset was comprised of 318 [50%] patients on standard therapy, as shown in **Figure 1**. All patients in the derivation dataset were randomized to six-months therapy duration. The validation datasets comprised of (i) 319 patients on standard therapy for six-months duration, and (ii) 1,287 patients randomized to the experimental arms [isoniazid or ethambutol] that had a four-months therapy duration. **Table 1** shows that the demographic and clinical characteristics were similar between the derivation data set and all validation data sets, which means that the data-partitioning step was executed successfully.

**Table 1.** Clinical features of patients in derivation and validation datasets.

| Variable | Total sample | Derivation dataset | Validation datasets |  |  | p-value |
| --- | --- | --- | --- | --- | --- | --- |
|  | n=1924 (%) | Standard therapy;<br>n=318 (%) | Standard therapy;<br>n=319 (%) | Ethambutol;<br>n=633 (%) | Isoniazid;<br>n=654 (%) |  |
| Age, years [mean (SD)] | 33.40 (12.16) | 33.10 (11.93) | 33.81 (12.40) | 33.88 (12.15) | 32.89 (12.17) | 0.442 |
| Sex, Female | 585 (30) | 91 (29) | 101 (32) | 188 (30) | 205 (31) | 0.767 |
| Male | 1339 (70) | 227 (71) | 218 (68) | 445 (70) | 449 (69) |  |
| **Race** |  |  |  |  |  | 0.663 |
| Black | 861 (45) | 149 (47) | 146 (46) | 289 (46) | 277 (42) |  |
| Asian | 586 (30) | 96 (30) | 96 (30) | 193 (30) | 201 (31) |  |
| Mixed | 451 (23) | 66 (21) | 74 (23) | 142 (22) | 169 (26) |  |
| Other | 26 (1) | 7 (2) | 3 (1) | 9 (1) | 7 (1) |  |
| **Country site** |  |  |  |  |  | 0.986 |
| China | 22(1) | 6 (2) | 2 (1) | 5 (1) | 9 (1) |  |
| India | 372 (19) | 58 (18) | 61 (19) | 126 (20) | 127 (19) |  |
| Kenya | 136 (7) | 26 (8) | 18 (6) | 43 (7) | 49 (7) |  |
| Mexico | 22 (1) | 7 (2) | 2 (1) | 8 (1) | 5 (1) |  |
| Malaysia | 69 (4) | 10 (3) | 13 (4) | 20 (3) | 26 (4) |  |
| Thailand | 119 (6) | 21 (7) | 19 (6) | 41 (6) | 38 (6) |  |
| Tanzania | 211 (11) | 37 (12) | 37 (12) | 73 (12) | 64 (10) |  |
| South Africa | 908 (47) | 142 (45) | 156 (49) | 297 (47) | 313 (48) |  |
| Zambia | 65 (3) | 11 (3) | 11 (3) | 20 (3) | 23 (4) |  |
| Sputum TTP in days<br>(SD) at start of therapy | 5.16 (1.21) | 5.26 (1.16) | 5.13 (1.26) | 5.15 (1.20) | 5.13 (1.21) | 0.420 |

### Time-to-extinction versus clinical trial-based outcome definitions

We then used the derivation dataset to determine if the time-to-extinction of the total *Mtb* sputum population for each patient have clinical relevance, especially given that TTP versus CFU/mL relationship could change with time during treatment. The number of patients deemed cured at different time intervals in the course of treatment obtained by counting the number of negative cultures/TTP as defined in the REMoxTB protocol versus those identified using our time-to-extinction model definitions [derived from CFUs calculated from TTPs] had a Spearman rank correlation of 1.0 [p=0.017]. Moreover, when we used Cohen’s kappa [κ] to assess agreement between individual pairs of either time-to-extinction versus standard clinical definitions, they were highly concordant [κ =0.65, p<0.001]. Furthermore, the Spearman rank correlation between *γ_f_* (fast slope) and 14-day extended-EBA [derived using linear regression] was 0.68 [p <0.001], which suggests that the extended-EBA mainly reflects the effect of treatment on *Mtb* in logarithmic-growth phase and not semidormant/NRP bacilli, as was assumed in the past [8].

### Predictors of outcome in derivation dataset

Classification and regression trees [CART] were used, to identify predictors of target outcome, defined as sputum microbial outcomes [cure at end of therapy, therapy failure or relapse], using potential predictors that included ALL the clinical and laboratory features, including ODE-model derived *γ*-slopes, for the tasks of classification and regression as input/independent variable. CART identified the *γ_s_* [semi-dormant/NRP kill] slope as the primary predictor [which had a variable importance score of 100%], followed by initial bacterial burden just prior to therapy commencement [which we termed *B* (0)], which had a variable importance score of 91.7%. This means that the initial TTP [*B*(0)] improved the primary predictor by an extra 91.7%. Notably, *γ_f_* was not ranked as a predictor using this agnostic machine learning method. CART performs its own cross-validation within the derivation dataset, in this case by randomly splitting the derivation dataset five times. With the cross-validation, the post-test validation area under the curve [AUC] in the same derivation set was >85%, demonstrating that *γ_s_* plus initial TTP [*B*(0)] will likely perform as good predictors in future datasets.

### Clustering-based approaches to identify biomarkers in derivation dataset

Clustering identified four distinct outcome groups based on individual trajectories versus time-to-extinction analysis in the 238 patients in the derivation dataset, as shown in **Figure 3**. These were, [1] a cure cluster of 80 (33.61%) patients, [2] a slow-cure cluster of 100 (42.02%), [3] a relapse cluster of 34 (14.28%) patients, and [5] a treatment failure cluster of 24 (10.08%) patients. The slow cure cluster identified by this unsupervised machine learning method denoted those patients who had delayed attainment of microbiologic cure at the end of six months therapy [failed therapy at the end of six months] but achieved relapse-free cure when standard therapy was continued beyond six months duration. These four clusters represented 238/318 [74.84%] of patients with less than 2 or more missing observations during follow up. The model explained these data well, as is shown in online **Figure S2, Figure S3**, and **Table S1**, while the corresponding summary statistics for each cluster are shown in **Table S2**.

**Figure 3.**
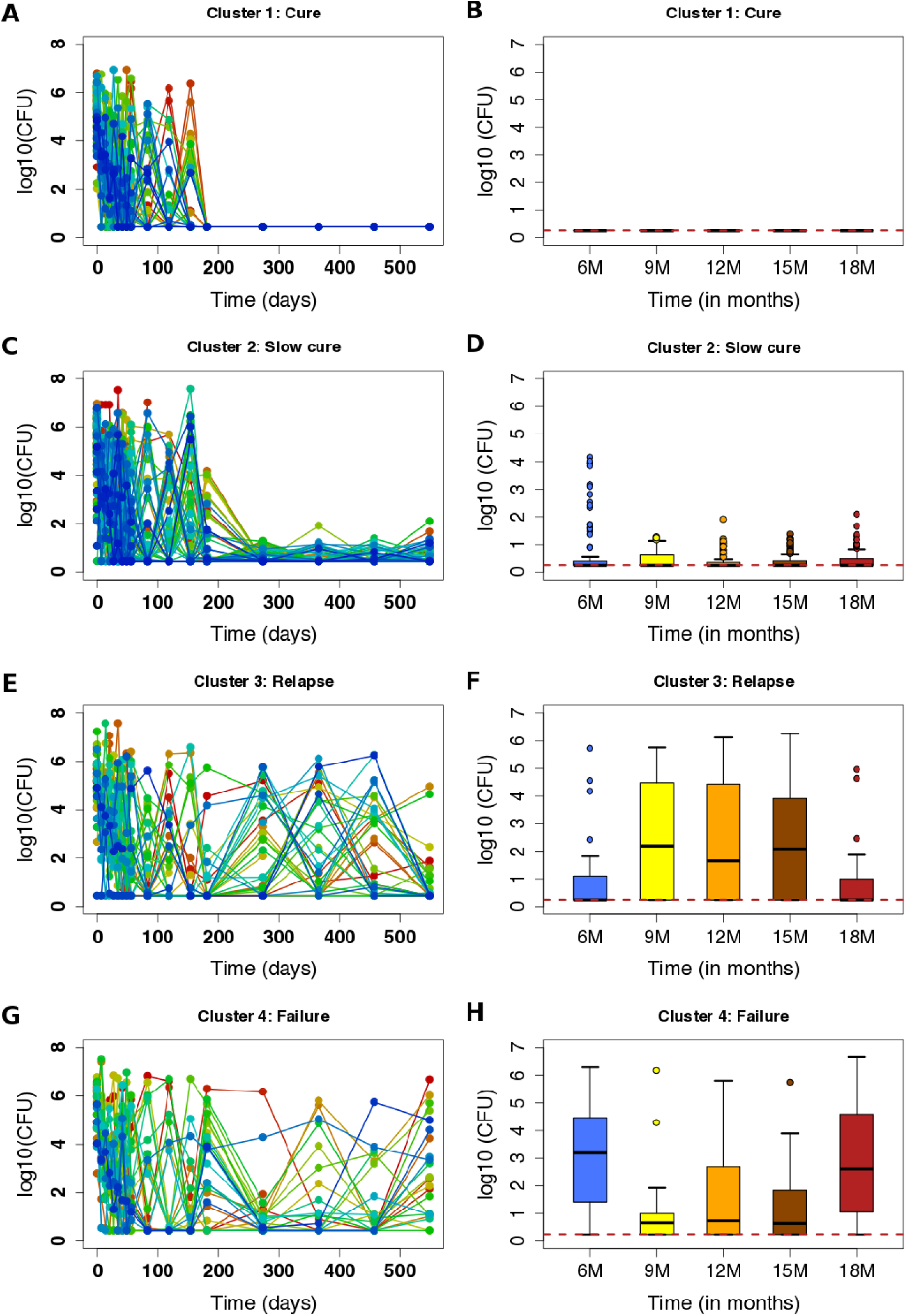
Clusters of treatment outcomes. Clusters of CFU/TTP trajectories of each individual patient are shown side by side with median log_10_ CFU/mL plus interquartile range, for the follow up periods of 6 to 18, months. (**A**) Cured patients’ trajectories and (**B**) summary of trajectories during follow-up. (**C**) Slow cure trajectories and (**D**) box-plots of CFUs after therapy completion. Relapse patterns (**E**) and the corresponding patterns during follow up (**F**). Failed treatment cluster (**G**) and follow-up period summarizing boxplots (H).

We used this clustering step to identify the minimum duration of data gathering that would give a γ-slope that could accurately predict cure or therapy failure or relapse. **Figure 4** shows the distribution of model derived *γ_s_* and *γ_f_* values, when these slopes were derived based on 8-weeks-derived TTP data [2-months], 4-months-derived TTP data, and 6-months-derived TTP data. The 8-week-, 4-months-, and 6-months-derived *γ_s_* and *γ_f_* values [shown in **Table S1**] versus outcomes were examined in pairwise comparisons using the Mann-Whitney-Wilcoxon test. **Figure 4** shows that the *γ_f_* values did not discriminate failures from cures, consistent with CART findings. However, *γ_s_*=0.15 or <0.1 log_10_ CFU/mL/day [modeling semi-dormant/NRP Mtb] were better at discriminating these outcomes. The slopes derived with 8-week-vs-4 months data differed in the misclassification of patients’ outcomes, the former misclassifying more relapses as cures and the latter misclassifying more cures as relapses. Nevertheless, as demonstrated by the statistical comparisons in **Figure 4I**, the 8-week derived TTP data *γ_s_* adequately diagnosed relapse versus other outcomes. In other words, *γ_s_* calculated using eight-week-derived TTP data is a good predictor of sterilizing effect up to 18-months after therapy cessation, and this eight-week data-derived slope thus measures sterilizing activity rate.

**Figure 4.**
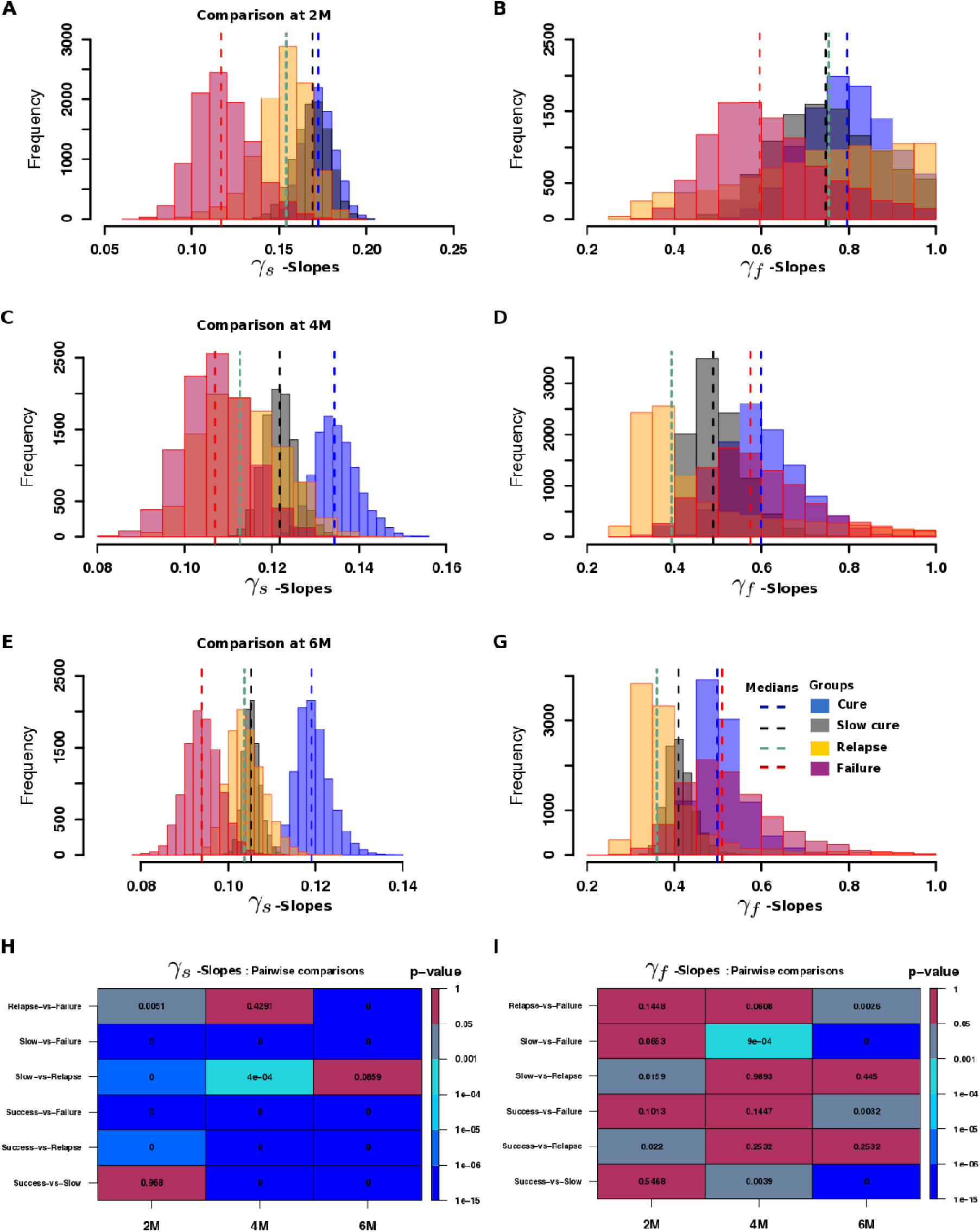
Distributions of model estimated kill rates/biomarkers. Distributions of *γ_s_* - and *γ_f_*-slopes based on 2-months, 4-months and 6-months accrued TTP data. (**A**) *γ_s_*-slopes based on 2 months TTP data, (**B**) *γ_s_*-slopes based on 4 months TTP data, and (**C**) *γ_s_*-slopes based on 6 months TTP data. (**D**) The magnitude of *γ_f_*-slopes at 2 months, (**E**) at 4 months, and (**F**) at the end of 6 months. (**H**) Pairwise analysis using the Mann-Whitney test for distributions of slopes based on 2, 4 and 6 months data versus each cluster (cure, slow-cure, relapse and failure) for the *γ_s_*-slopes shows p-values in each cell that were significant even with 2-months of data, except with cure versus slow cure group. (**I**). Pairwise statistical difference analysis for *γ_f_*, found that few p-values were significant, and even those had an inconsistent pattern.

Subsequently, all *γ_s_* discussed herein were those identified using the first eight-weeks-derived data.

### Monte Carlo Simulations to identify biomarker thresholds in indeterminate outcome zones

Given the misclassification of relapses as cures by the eight-week TTP-derived *γ_s_*, and *γ_s_* thresholds in the indeterminate outcomes region [i.e, overlap of relapse versus slow cures], we utilized Monte Carlo Simulations [MCS] of time-to-extinction in tandem with CART to further discriminate *γ_s_* cut-off values in indeterminate outcome zones, with results shown in **Table S3**, **Figure S4** and **Figure S5**. Cure was clearly delineated by *γ_s_* >0.15, therapy failure by *γ_s_*<0.1 plus initial bacterial burden *B*(0)>5.6 log_10_ CFU/mL [TTP=5.49 days], and relapse delineated from cure by *γ_s_*<0.13. **Figure S4C-D** shows the CART-derived biomarker thresholds based on the simulation for predicting treatment outcomes after either 4-months or 6-months therapy duration. Patients with initial bacteria burden *B*(0)>4.5 log_10_ CFU/mL [TTP=8.11 days], and *γ_s_*-slopes between 0.1 and 0.15 had >55% chance of failing treatment at 6 months [**Fig. S4C**]. However, for a four-month therapy duration regimen, patients with *B*(0)>5.4 log_10_ CFU/mL [TTP=5.93] and *γ_s_* between 0.09 and 0.14 had a >65% chance of failing treatment. **Figure S4** also shows that in order to achieve cure/bacillary population extinction within 2 months of treatment, then *γ_s_*≥0.15 [-3.90 TTP per day] would be required, while patients with *γ_s_*≤ 0.1 [-2.60 TTP per day] would fail. Patients on standard therapy with *B*(0)>5.6 log_10_ CFU/mL [TTP=5.49] with *γ_s_* <0.13 would relapse.

### Creation of *γ_s_*-based rule to predict relapse for different therapy durations from derivation dataset

In the final derivation step, we established a diagnostic rule for the relationship between *γ_s_*-slopes and the outcomes, using Latin hypercube sampling for sensitivity analyses, with results shown in **Figure 5**. **Figure 5A-D** shows that increasing or reducing the *γ_s_* [i.e., speed of kill of slow-replicating bacteria] changes the time-to-extinction and therefore the required minimum duration of therapy. As an example, the six-months therapy duration would need to be extended to eight-months duration [i.e., slow-cure] in patients with high bacterial burden when *γ_s_* is reduced from 0.148 to 0.131 and extended to 9-months when its reduced to 0.125 [**Figure. 5B]**. However, for patients in the medium and low CFU load categories, lower slopes can still achieve cure within 6 months [**Figure. 5C-D]**. On the other hand, to reduce treatment duration to four-months *γ_s_* should increase to 0.183, and in order to reduce therapy duration to two-months *γ_s_* should increase to 0.286 [**Figure. 5B]**. The relationship betweeny and initial TTP versus minimum duration of therapy is shown in **Figure. 5E-F**, and is non-linear function. From this, we calculated the target *γ_s_* to achieve cure [extinction of bacterial population] with one-month therapy duration, shown in **Figure. 5E-F**. This establishes a diagnostic rule between *γ_s_* versus minimum treatment duration for relapse-free cure for different initial *Mtb* burdens. After this step, the derivation work was completed, and the derivation dataset patients excluded from subsequent validation studies.

**Figure 5.**
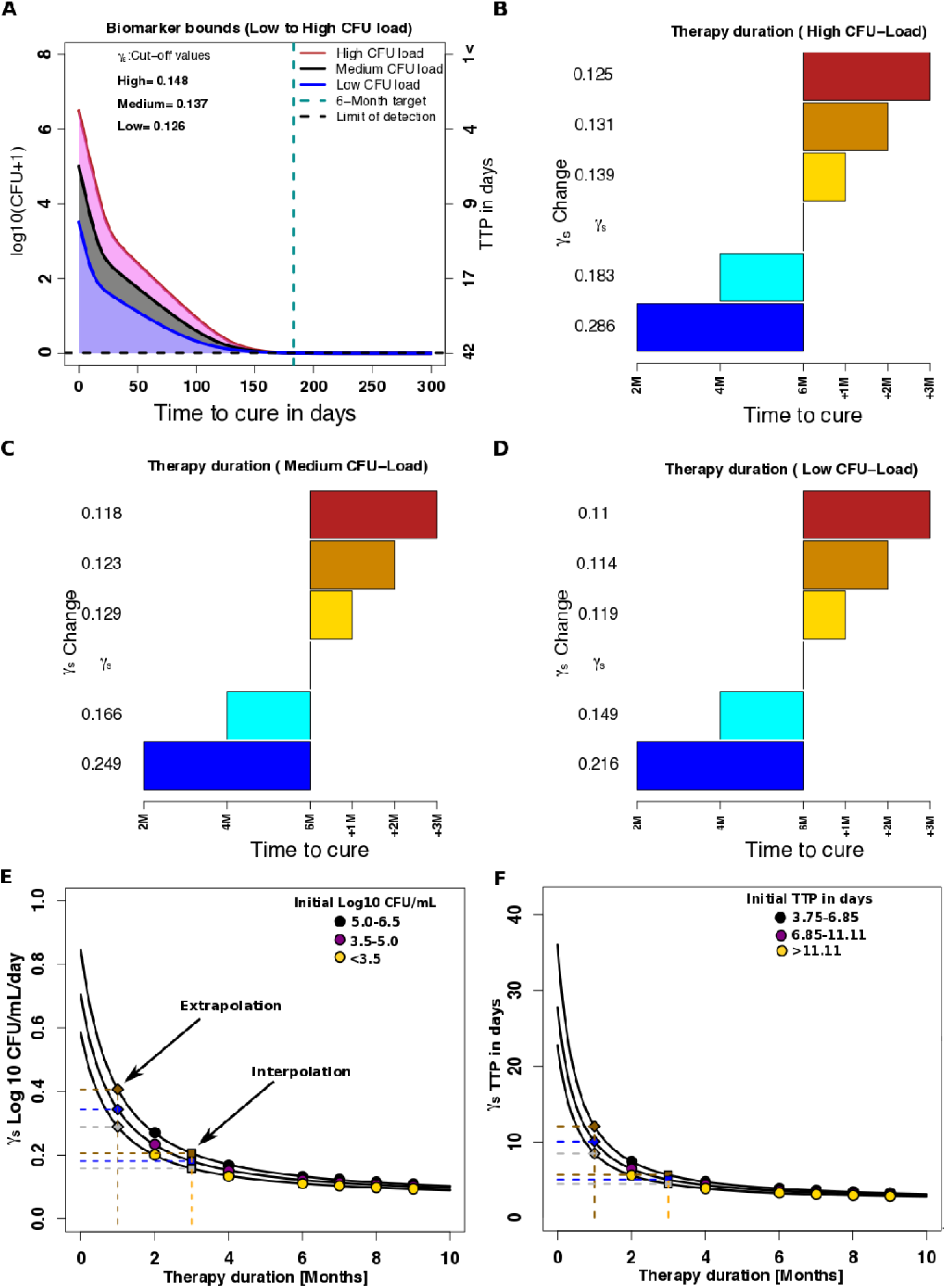
Sensitivity analyses and rule-making of *γ_s_* slopes versus time-to-cure. (**A**) Shown are *γ_s_* slopes required to achieve cure within 6 months for patients with high bacterial burden compared to those with medium and lower bacterial burdens. (**B-D**) The *γ_s_* slopes required to achieve cure at 2, 4 and 6 months duration or for delayed cure of an additional 1 to 3 months beyond month 6 [i.e, 6 months +1, or +2, or +3 months], are shown for patient starting with high (**B**), medium (**C**), and low (**D**) *Mtb* burdens. (**E**). Magnitudes of slopes for therapy duration of only 1 and 3 months (for high, medium and low *Mtb* burden) could be extrapolated and interpolated in log_10_ CFU/mL/day as (0.42, 0.36, and 0.30) and (0.22, 0.20, and 0.17), respectively, based on the relationships between slope and duration of therapy (r^2^>0.999). (**F**) Magnitudes of slopes for therapy duration of 1 and 3 months are extrapolated and interpolated TTP-slope as (12.08, 10.08 and 8.49) and (5.67, 5.06 and 4.48), respectively, based on the relationships between TTP-derived slope and duration of therapy (r^2^>0.999).

### Performance of *γ_s_*based rules in forecasting outcomes for 6 months therapy duration

Next, we calculated the accuracy of how well our diagnostic rule performed in the six months therapy duration validation datasets, using the clinical and microbial treatment outcomes defined by the REMoxTB trial protocol. Treatment outcome calls could be made in 218 of the 319 patients who also had more than 4 data points within eight-weeks to give statistically robust estimates of the bacteria kill slopes: 169/218 (74.31%) achieved relapse-free cure, 137/218 (16.97%) had therapy failure at the end of treatment, and 19/218 (8.72%) relapsed after initially looking like cure at the end of therapy. The accuracy of the *γ_s_*-based rules are compared to the extended-EBA and two months sputum conversion in **Table 2**, together with the relative risk [RR] of failure when each biomarker was positive versus not-positive [numbers in each cell shown in **Table S4**]. **Table 2** shows that the extended-EBA had a sensitivity of 14% and specificity of 92% in identifying failure from cure without relapse and the RR 95% confidence interval crossed 1 [p=0.205]; the number needed to diagnose [NND] failure/relapse was 15.27. Similarly, two-months sputum conversion had a sensitivity of 33% and specificity of 71%, RR was statistically 1, and NND was 21.41. On the other hand, the eight-weeks-data derived γ*_s_* combined with the initial TTP at treatment commencement had a sensitivity of 92% and specificity of 86% in identifying failure from relapse-free cure, the RR of failure when this biomarker was positive versus not positive was 28 [**Table 2 and Table S4**], while NND was 1.29. Failures either arise as therapy failure or relapse; **Table 2** shows the sensitivities for these different biomarkers in predicting relapses from treatment failures. The slope decision rule based on *γ_s_*>0.15 has a sensitivity of 92% and a specificity of 89% in predicting relapses from failures. Thus, the biomarkers we derived were highly specific at identifying relapse-free cure, therapy failure, and relapse.

TTP (0) is the corresponding TTP in days for *B* (0): A dash means no cut-off value evaluated. The thresholds for predicting relapses-vs-cure are multiple steps however, are with the 0.1 to 0.15 indeterminate regions of the slow slope cut-offs for screening cures and failures.

**Table 2.** Biomarker threshold values, sensitivity and specificity scores, and risk of failure, with 95% confidence intervals.

| Biomarker using log<sub>10</sub> CFU/mL | Sensitivity | Specificity | Accuracy | Relative risk of failure with positive biomarker |
| --- | --- | --- | --- | --- |
| **Six-months therapy duration** |  |  |  |  |
| Extended EBA: Failure vs Cure | 0.14 (0.05-0.30) | 0.92 (0.87-0.96) | 0.79 (0.73-0.84) | 1.71 (0.73-3.48) |
| 2 months smears/culture: Failure vs Cure | 0.33 (0.17-0.53) | 0.71 (0.64-0.78) | 0.65 (0.58-0.72) | 1.20 (0.60-2.34) |
| $\gamma_s^1$ slope (<0.1): Failure vs Cure | 0.57 (0.39-0.73) | 0.95 (0.91-0.98) | 0.89 (0.84-0.93) | |
| $\gamma_s^2$ slope (>0.15): Failure vs Cure | 0.91 (0.78-0.98) | 0.86 (0.79-0.90) | 0.87 (0.81-0.91) | |
| Slow slope plus $B(0)$: Failure vs Cure<br>$\gamma_s^1 \gamma_s^2, B(0)$ | 0.76 (0.59-0.88) | 0.89 (0.83-0.93) | 0.87 (0.81-0.91) | 29.84 (10.20-89.07) |
| Slow slope plus $B(0)$: Failure-vs-Relapse<br>$\gamma_s^1 \gamma_s^2, B(0)$ | 0.92 (0.78-0.98) | 0.89 (0.67-0.99) | 0.91 (0.80-0.97) | 20.40(7.17-58.08) |
| **Four months therapy duration** |  |  |  |  |
| **Isoniazid arm** |  |  |  |  |
| Extended EBA: Failure vs Cure | 0.10(0.04-0.21) | 0.95(.93-0.98) | 0.84(0.80,0.87) | 2.14 (0.99-3.99) |
| $\gamma_s^1 \gamma_s^2, B(0)$: Failure vs Cure | 0.81 (0.70-0.90) | 0.87 (0.83-0.90) | 0.86 (0.83-0.89) | 14.51 (8.33-25.41) |
| $\gamma_s^1 \gamma_s^2, B(0)$: Failure vs Relapse | 0.75(0.58-0.87) | 0.60(0.51-0.69) | 0.64(0.56-0.71) | 3.15(1.65-6.01) |
| **Ethambutol arm** |  |  |  |  |
| Extended EBA: Failure vs Cure | 0.10(0.05-0.19) | 0.94(0.92-0.96) | 0.79(0.76-0.83) | 1.66 (0.89-2.81) |
| $\gamma_s^1 \gamma_s^2, B(0)$: Failure vs Cure | 0.70 (0.60-0.79) | 0.71 (0.67-0.75) | 0.71 (0.67-0.75) | 4.10 (2.78-6.08) |
| $\gamma_s^1 \gamma_s^2, B(0)$: Failure vs Relapse | 0.70(0.59-0.79) | 0.65(0.55-0.74) | 0.68(0.60-0.74) | 2.07(1.50-2.87) |

TTP (0) is the corresponding TTP in days for $B(0)$ : A dash means no cut-off value evaluated. The thresholds for predicting relapses-vs-cure are multiple steps however, are with the 0.1 to 0.15 indeterminate regions of the slow slope cut-offs for screening cures and failures.

### Performance of *γ_s_*-based rules in forecasting 4-months therapy duration outcomes

Next, we tested the accuracy of the diagnostic rule for four-months therapy duration in the validation datasets comprised of the REMoxTB trial experimental arm patients. In the arm in which isoniazid was replaced by moxifloxacin and therapy administered for four-months (n=655), 530 patients had enough TTP data in the first 8 weeks to calculate slopes. In this dataset, 369/530 (69.62%) patients achieved cure, 40 (7.55%) patients had therapy failure at the end of 4 months of therapy, and 121 patients (22.83%) relapsed. **Table 2** shows that the γ*_s_>*0.15 had a sensitivity of 81% and specificity of 89% for relapse-free cure versus failure, and among the failures had a sensitivity of 75% and specificity of 60% for separating relapse from therapy failure. The relative risk of failure in patients with positive slope-based biomarker versus negative biomarker was approximately 15 [**Table 2 and Table S4**]; the NND was 1.47. The 2-month sputum conversion was not designed for 4 months therapy duration regimens, and is not shown, while the extended-EBA which is used to triage shorter duration regimens is shown; the NND was 16.69.

In the arm in which ethambutol was replaced by moxifloxacin (n=633), 533 patients had enough data to calculate 8-week slopes. In this dataset 385 (72.23%) of patients achieved cure, 46 (8.63%) had therapy failure, while 102 relapse (19.4%). The sensitivity of the extended EBA was only 10%, and the NND was 18.73. The sensitivity of *γ_s_*-based slopes was 70% and the specificity 71% for cure versus therapy failure, while the sensitivity was 70% and specificity 65% for picking relapse versus therapy failure. The NND was 1.89.

In order to summate, we calculated an overall value of the relative risk of failure when our *B*(0) and *γ_s_*-based slope predicted poor outcome for a specified duration of therapy [using 6-months and 4-month duration data combined]. Among patients with positive biomarker for specified therapy duration, 159/205 [78%] failed therapy compared to 218/1072 [20%] in whom the biomarker was negative. The RR of failure with the rule was 8.25 [95% CI: 6.09-11.20]; p<0.0001. In terms of cure only 4% of entire validation dataset cohort of patients achieved relapse-free cure when our rule was positive while 67% achieved cure when it was negative.

## DISCUSSION

First, we found that the *γ_s_* [slow replicating] slope is a good surrogate of sterilizing activity, based on ability to predict relapse. Conversely, the extended EBA had a sensitivity of 14% for predicting outcomes at 6 months and beyond, and a poor accuracy. The extended EBA is effectively two-weeks accrued data; the poor sensitivity means that the total time for which the bacterial kill data is collected is too short to accurately capture sterilizing activity slopes. Indeed, the poor sensitivity of *γ_f_*-slope-based metric means that most regimens with good sterilizing effect could be thrown away [too many false negatives for sterilizing activity] in regimen selection for sterilizing activity. Similarly, the 2-month sputum conversion had a sensitivity of 33% and specificity of 71%. These commonly used clinical indices gave us an opportunity to externally validate our modeling approach. In this case, the last major meta-analyses on 2-month cultures as a predictor of long-term outcome in TB performed by Horne et al in 2010 identified a sensitivity of 40% [95% CI, 25-56%] and specificity of 85% [95% CI, 77%-91%], which was confirmed in subsequent studies [14, 41, 42]. Thus, our modeling findings are consistent with results of these major meta-analyses. This means that our 8-weeks-derived *γ_s_* slope plus initial bacterial burden, which had a sensitivity of 92% and specificity of 86% for 6 months therapy duration regimens, would perform better than the 2-month sputum conversion. In addition, our *γ_s_* slope can predict outcomes at shorter therapy durations than 6 months such as 4-months duration; the relative risk of therapy failure among patients with positive biomarker for specified therapy duration was >8.0 Thus the *γ_s_*-slope based on the first 8-weeks TTP data is a good response biomarker for sterilizing activity, even for therapy duration less than standard short course chemotherapy.

The *γ_s_*-slope, which we will henceforth term the “sterilizing activity rate”, fulfills the BEST criteria and definition of a monitoring biomarker in the category of a pharmacodynamic/response biomarker, in a similar fashion to HIV and hepatitis C viral load biomarker, and could play the same role in TB therapeutics and clinical trials [23, 43–45]. According to BEST criteria, a pharmacodynamic/response biomarker provides early evidence [in this case 8-weeks] that a treatment might have an effect on a later pharmacologic clinical endpoint [in this case relapse at 2 years]. In the case of HIV treatment trials, identification of viral load as a surrogate of efficacy in 1995 dramatically cut the duration and costs of clinical trials, while avoiding use of potentially catastrophic clinical endpoints such as therapy failure and death [43]. For TB, we propose identification and ranking of regimens using preclinical models that can accurately translate the sterilizing activity rate to patients [24, 46]. The regimens so derived, including optimal doses, and the translated sterilizing activity rate will provide good priors for the design of 8-week clinical trials for novel regimens versus standard therapy, with weekly TTP as the main output and drug pharmacokinetics as a secondary outcome. The sterilizing effect rate [*γ_s_*-slope], initial TTP, and trajectories can then be used to estimate therapy duration for the novel regimens and determine if indeed the new regimens can shorten TB treatment prior to performance of phase III studies. The 8-weeks TTP-data derived slopes can be used to compute a lower and more accurate patient sample sizes required to power the phase III trials, given the good accuracy in forecasting relapse. As an example, the number needed to diagnose [NND] failure and relapse of <2, when compared to ~20 for extended EBA and 5-6 for 2-months therapy, gives a more straightforward insight into the relative number of patients tested in each arm by different biomarkers. Moreover, since the predictive value of the sterilizing activity rates on relapse or cure or therapy failure is independent of the regimen the slopes can be used in clinical trials of MDR-TB and for “pan-susceptible” TB regimens, indeed for any TB regimen.

As regards to clinical practice, our findings add to the recent discovery that initial *Mtb* burden can be used to determine patients who can benefit from 4-month duration therapy [47]. Here, we found that the sterilizing activity rate was ranked higher than initial bacterial burden. To put this is context, the risk of development of AIDS and death in patients whose HIV viral load did not reach undetectable within first 12 months was 2.40-fold compared to those who had, and a <75% reduction in viral load had a RR of 2.27-fold for poor outcomes [48–50]. Patients in whom the *γ_s_*-slope-based rule was positive for different durations of therapy had a an 8.25-fold higher risk of failure, which is better performance than this commonly used HIV test used to individualize therapy. Thus, our findings could also be used to individualize therapy, in place of two-month smears/cultures currently recommended in routine care in TB programs worldwide. First, if these patients with potentially higher rates of therapy failure and relapse were identified during the first eight weeks of therapy, then interventions such as dose increases or switching therapy regimens could be made [37]. Second, the sterilizing effect rate *[γ_s_* slopes] could also be used by TB programs to identify patients who could be cured with specific shorter therapy durations of either 2, 3 or 4 months, on any regimen. Alternatively, they could be used to identify how long therapy duration should be extended beyond 6 months, thereby individualizing therapy duration, in patients with sputum *γ_s_* slopes that predict the slow cure clusters. Since many TB programs across the world already employ liquid culture systems that generate TTP, it means that the biomarker we propose would come at no extra cost to those TB programs. Computation of the slope could easily be implemented on a computer [or on a phone with specifically designed app].

Our study has some limitations. First, it could be argued that our findings are specific to the dataset we analyzed. However, the machine-learning cross-validation procedures we used are scored on how well predictors will perform on an entirely independent dataset in the future. Nevertheless, the accuracy of the biomarkers will still need to be further confirmed in other large datasets in a range of clinical contexts and with different regimens. Further, this approach can be adapted for other non-tuberculosis bacterial infections. Second, calculation of slopes is relatively complex. However, software can easily be written to automate this, as we have attempted elsewhere. Finally, not all patients who do not reach bacterial population extinction will fail therapy or relapse. This means that our approach may lead to over treating of these patients who would otherwise be cured. Examination of our proposed biomarkers with other tests such as radiological findings and therapeutic drug monitoring could reduce the number of over treated patients and are subject to ongoing analyses. However, even with these limitations, the early TTP-based biomarkers that we identified as predicting long-term clinical outcomes such as relapse for different therapy durations, have sensitivities and specificities that are higher than currently employed methods.

## Data Availability

Data will made available upon request to the authors

## ACKNOWLEDGEMENTS

We thank Dr. Stephen H Gillepsie and Dr. Patrick Phillips for help with insightful comments on the manuscript, and general advice about the REMoxTB study design and data.

## AUTHOR CONTRIBUTORS

GM performed the mathematical modeling, interpreted the data and wrote the first draft of the manuscript; JGP performed sample size calculations, CART and statistical analyses, and designed the sample size calculator; TG oversaw the conduct of the study, led the data interpretation, application of criteria for biomarkers, and the clinical meaning. All three authors wrote the manuscript, revised it, and all approved the final submitted version.

## Supplementary Material

**Table S1.**
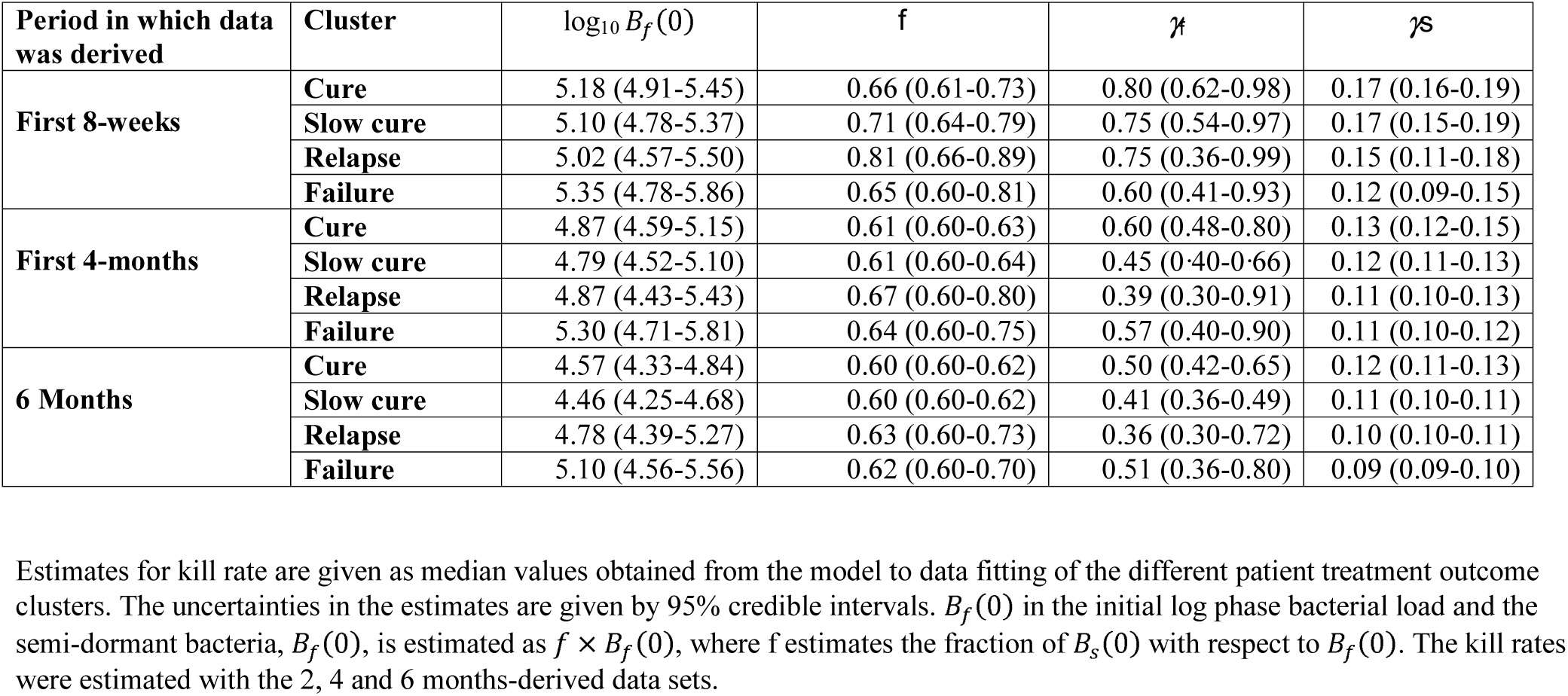
Estimated kill rates and initial bacterial load by cluster versus period of data derivation.

**Table S2.** Cluster summary for bacterial burden.

| **Cluster** | **6-Months** | **9-Months** | **12-Months** | **15-Months** | **18-Months** |
| --- | --- | --- | --- | --- | --- |
| **Cure** | 0.24 | 0.24 | 0.24 | 0.24 | 0.24 |
| **Slow cure** | 0.24 (0.24-0.40) | 0.24 (0.24-0.43) | 0.24 (0.24-0.40) | 0.24 (0.24-0.39) | 0.24 (0.24-0.48) |
| **Relapse** | 0.24 (0.24-1.11) | 2.20 (0.31-4.40) | 1.65 (0.24-4.43) | 2.10 (0.24-3.88) | 0.24 (0.24-1.00) |
| **Failure** | 3.19 (1.42-4.46) | 0.66 (0.24-1.00) | 0.73 (0.24-2.53) | 0.64 (0.24-1.84) | 2.61 (1.06-4.50) |

Summary estimates for different clusters during follow up period. Median estimates are given and the 1<sup>st</sup> and 3<sup>rd</sup> quartile values are given in brackets. The value 0.24 log10 CFU represent the limit of detection that corresponds to 42 days for TTP MGIT readouts.

**Table S3.** Two month-derived *γ_s_* magnitude cut-off values and initial bacteria burden.

| **Conditional cure regions** | $\gamma_s$ cut-off [log<sub>10</sub> CFU/mL/day] | $\gamma_s$ cut-off [TTP day] | **Initial bacterial [TTP/day]** |
| --- | --- | --- | --- |
| 6 months outcomes | $0.1 < \gamma_s < 0.15$ | $-3.9 < \gamma_s < -2.6$ | -<8.11 |
| 4 months outcomes | $0.09 < \gamma_s < 0.14$ | $-2.86 < \gamma_s < -2.34$ | -<5.93 |
| **Failure regions** |  |  |  |
| 6 months outcomes | <0.1 | < -2.60 | - |
| 4 months outcomes | <0.1 | < -2.60 | - |
| **High chance of relapse regions** |  |  |  |
| 4 months and 6 months outcomes | $0.1 < \gamma_s < 0.15$ | $-3.9 < \gamma_s < -2.6$ | <-5.49 |

**Table S4.** Contingency tables of outcomes versus different biomarkers.

|  | **8week-Biomarker** |  | **Extended EBA** |  | **2 month smears** |  |
| --- | --- | --- | --- | --- | --- | --- |
|  | **Positive** | **Negative** | **Positive** | **Negative** | **Positive** | **Negative** |
| **6 months therapy duration** |  |  |  |  |  |  |
| Cure | 3 | 155 | 30 | 155 | 20 | 117 |
| Failure | 34 | 26 | 5 | 13 | 10 | 47 |
| **4 months therapy duration- isoniazid arm** |  |  |  |  |  |  |
| Cure | 13 | 361 | 53 | 344 | 36 | 299 |
| Failure | 56 | 55 | 6 | 15 | 20 | 47 |
| **4 months therapy duration- ethambutol arm** |  |  |  |  |  |  |
| Cure | 30 | 338 | 77 | 378 | 61 | 334 |
| Failure | 69 | 137 | 9 | 23 | 25 | 76 |

**Figure S1.**
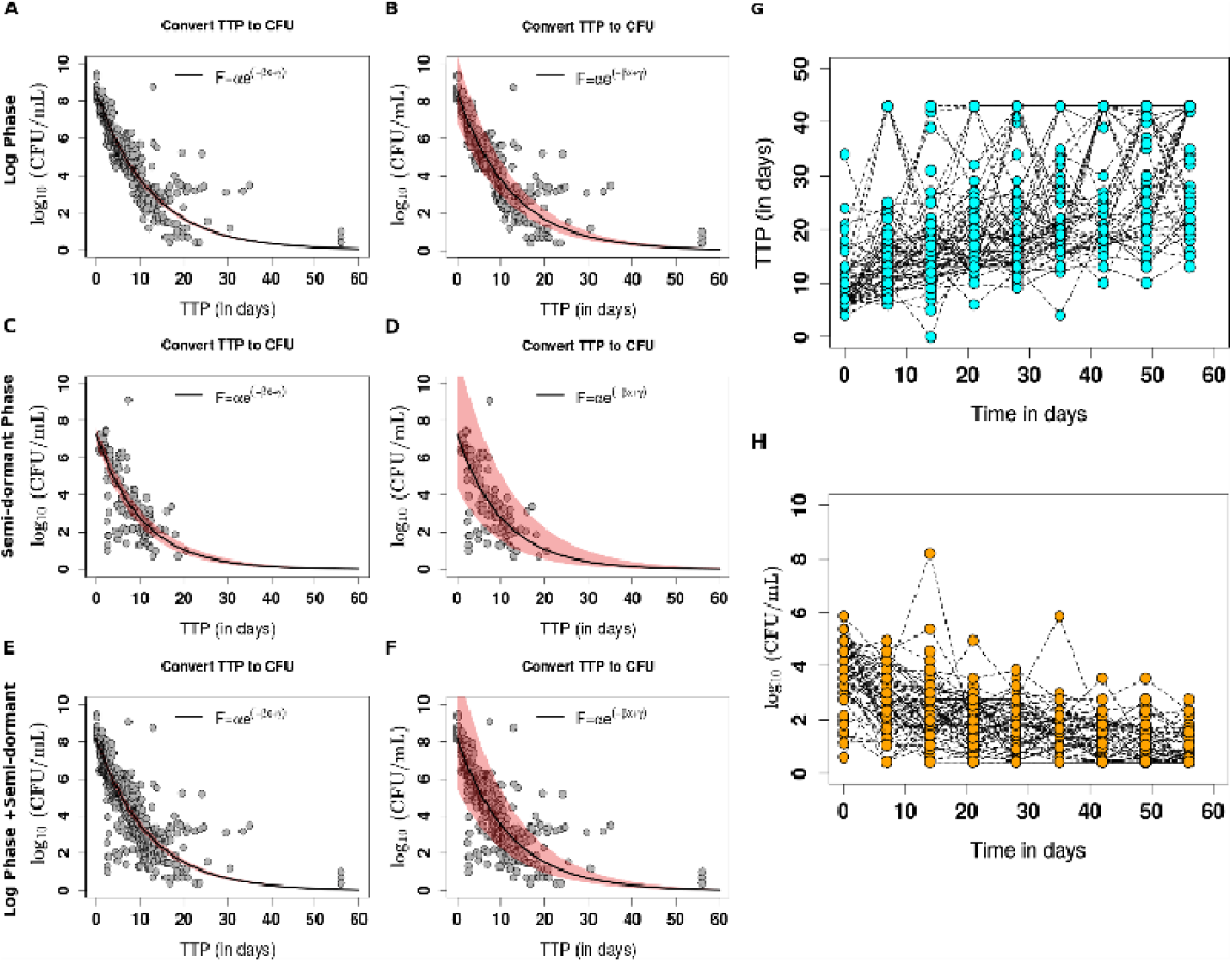
Converting TTP to CFU and vise-versa. Illustrating conversion of serial TTP values from patients to corresponding CFU values. Panels **A** showing model function correlating TTP to CFU, here the model has two parameters to the estimated, in **B** the model with three parameters has wider uncertainty. While conversion for bacteria in log phase is shown in **A** and **B**, conversion for slow (non replicating) replicating bacteria is shown in **C** and **D**, then in panels **E** and **F**, combined subpopulations are shown. Panel **G** shows TTP patient clinical data and the corresponding converted CFU are shown in **H** at start of therapy [Day 0] and every 7 days till day 56.

**Figure S2.**
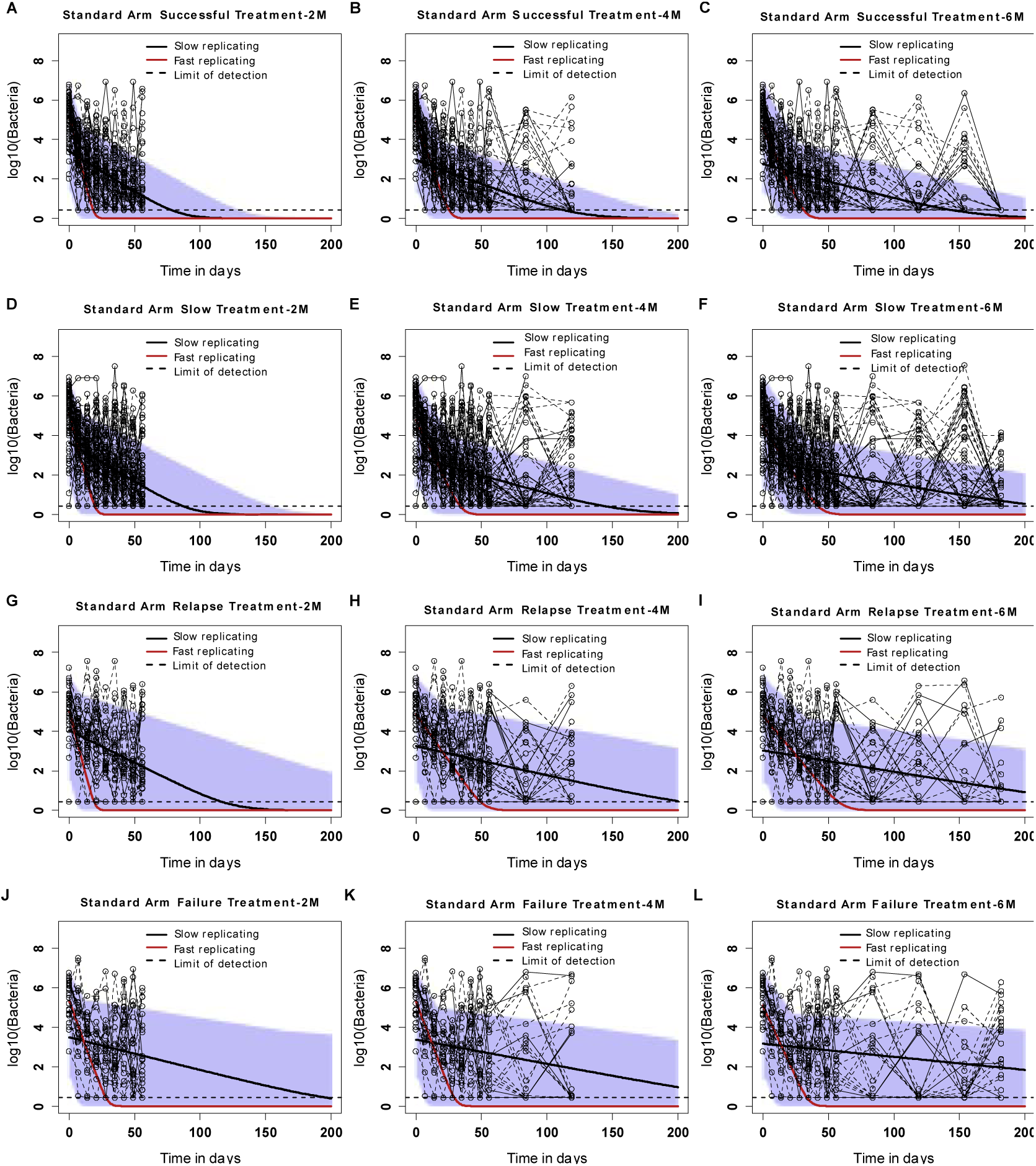
Model fitting to accrued data at 8-weeks, 4-months and 6-months time points. Shown are model fitting to the accrued cure data cluster at 8-weeks/2M (**A**), 4M (**B**) and 6M (**C**), respectively. Model fitting to the slow successful cure cluster data are shown in **D** (at 2M), **E** (at 4M) and **F** (at 6M). Relapsing disease data patterns are given in **G**, **H**, and **I** and while treatment failure are shown in **J**, **K** and **L** at 2M, 4M and 6M, respectively. The gold dots represent the observations, the solid lines are the model predictions and the shaded regions represent the 95% credible intervals.

**Figure S3.**
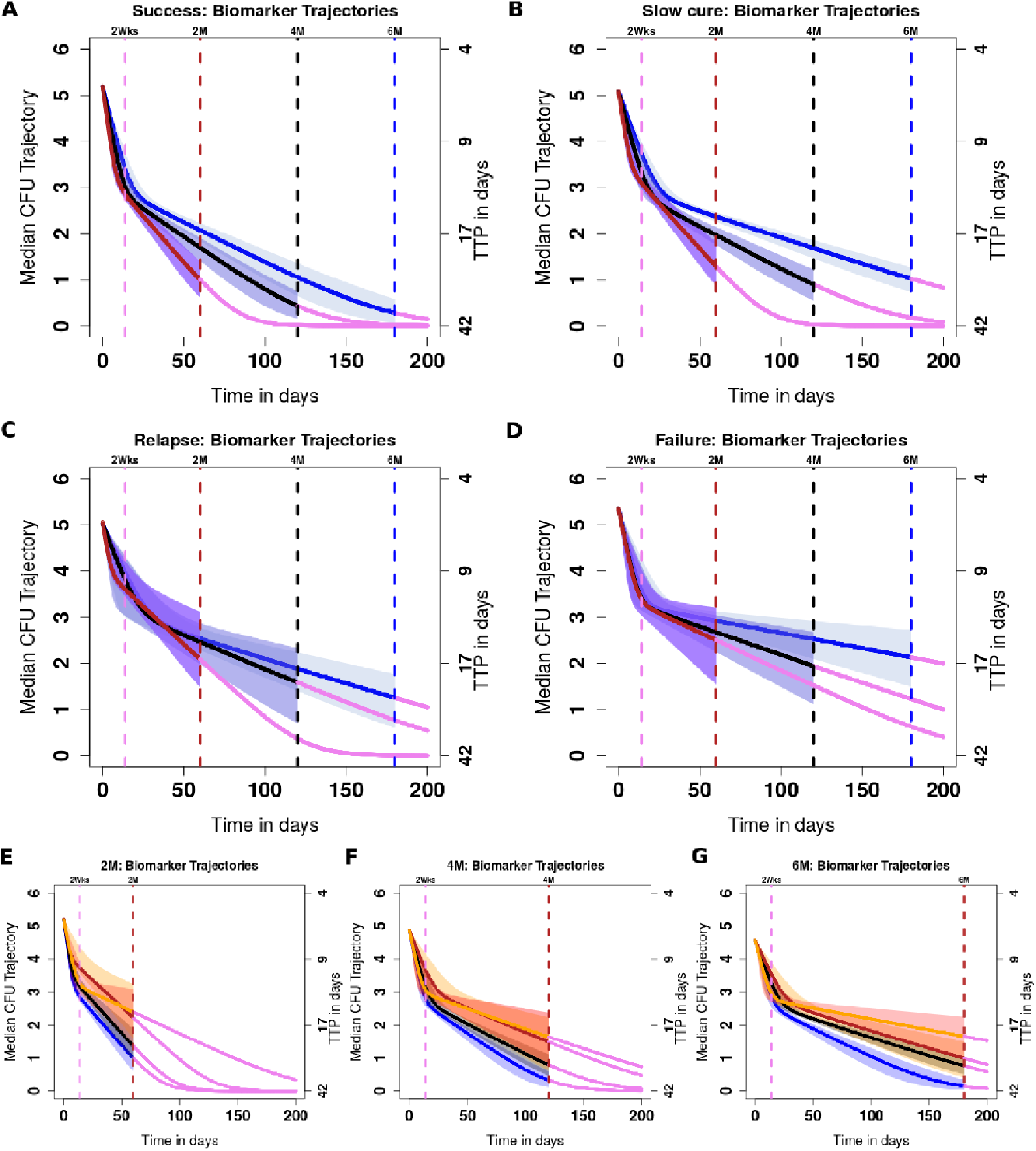
Evolution of treatment outcome biomarkers. (**A**) The evolution of biomarkers patterns that identifies cured patients at 2M, 4M and 6M, respectively. Summary trajectories for patients in the slow successful treatment cluster (**B**), relapsing cases (**C**) and treatment failure cluster (**D**) demonstrate how these patterns diverge. Further comparisons of all biomarkers for each cluster and their respective 95% credible intervals at 2M (**E**), at 4M (**F**) and at 6M (**G**) end points are shown.

**Figure S4.**
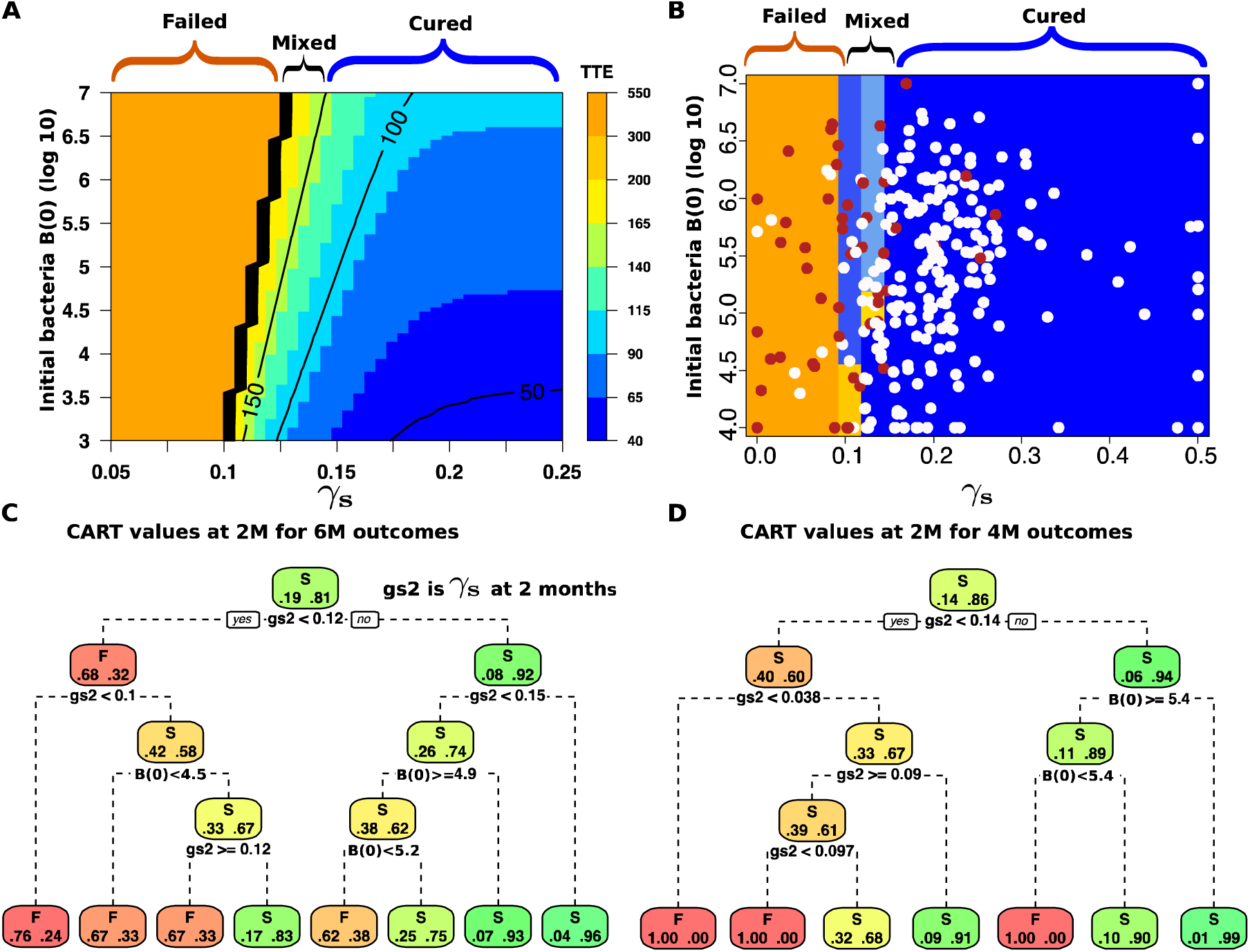
Biomarker characterization in indeterminate data zones. Thresholds for the biomarkers that enable treatment outcome predictions at 2, 4 and 6-months are shown. (**A**) Simulation-based derivation of the biomarker thresholds and how they vary with initial patient bacteria burden, *B*(0). (**B**) A combination of *γ_s_*-slopes and *B*(0) in classifying outcomes classification regression trees (CART). In **C** and **D**, shown are the biomarker breakpoints predicted with CART using 6M and 4M treatment outcomes, respectively. In **C**), failures at 6 months are predicted with a *γ_s_*-slope that is less than 0.1. In **D**), the 4-month failure threshold is predicted to be 0.1(0.097), while 0.14 predicts cures. Both in C and D, the zone between *γ_s_*-slope of 0.1 and 0.15 has high misclassification. The mixed regions in **A** and **B** demonstrate conditional outcomes dependent on initial bacterial burden and *γ_s_* rates (**Figure S4**).

**Figure S5.**
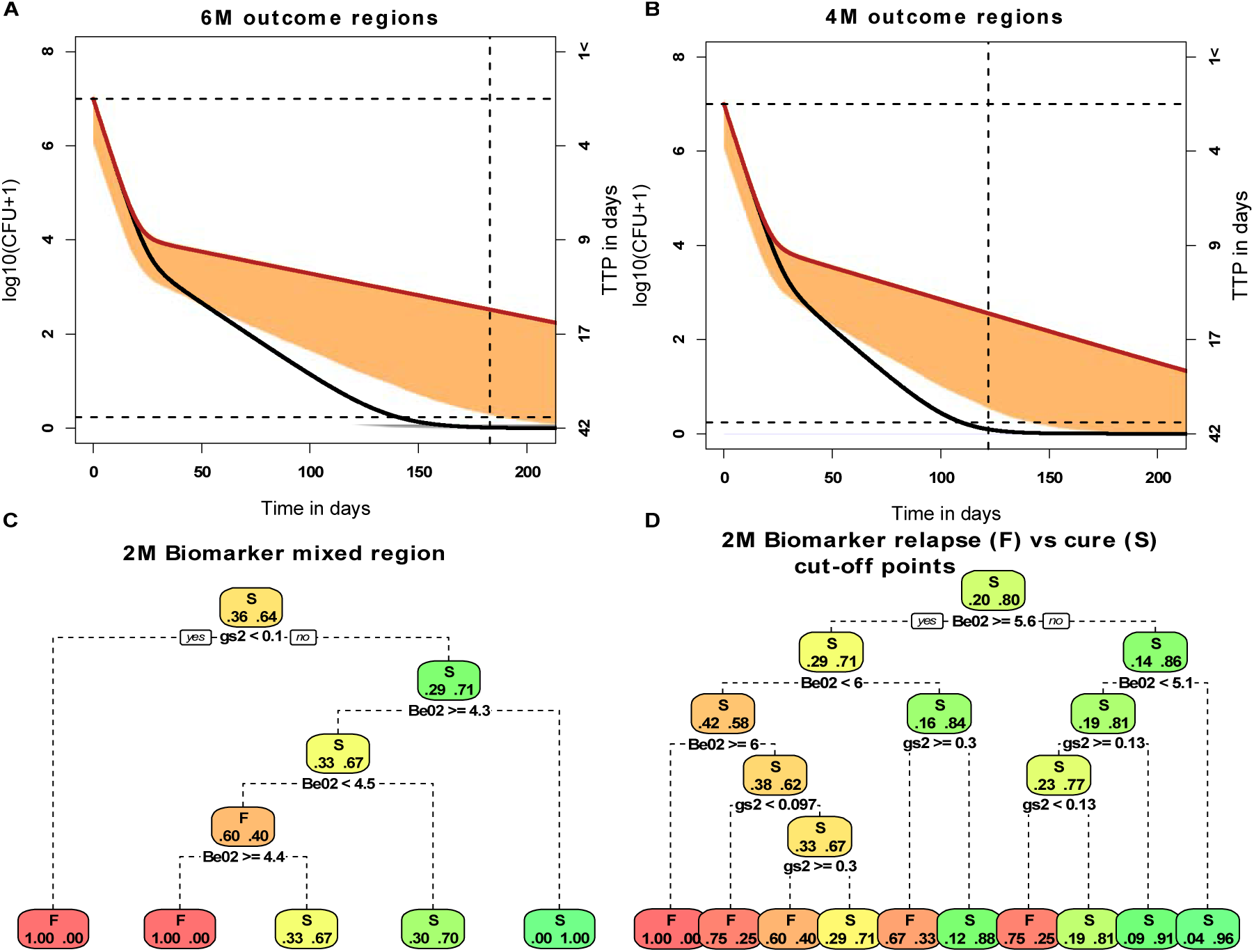
Thresholds for regions with mixed outcomes. *γ_s_* values greater than 0.125 cure patients with initial bacteria burden between 4-6 log 10CFU/mL and values below 0.125 will result in failure using simulations. (**B)** illustrates the overlap between the initial bacteria and the slopes for the 4-month outcomes. In **A** and **B** regions of failure are shaded orange and the cure region has the grey shading). (**C)** CART derived cut-off values for 6 months outcomes. **(D)** CART derived predictor (initial burden and *γ_s_*-slope) cut-off to delineate relapses from cure.

